# Parameters for assessment of talar neck geometry: a protocol for scoping literature review

**DOI:** 10.1101/2020.09.28.20203562

**Authors:** Siddhartha Sharma, Karan Jindal, Sandeep Patel, Sharad Prabhakar, Mandeep S. Dhillon

## Abstract

**Background:** Understanding the three-dimensional anatomy of the talar neck is essential in assessing reduction of talar neck fractures, as well as in planning surgical correction for talar malunions. However, the geometrical parameters that describe this anatomy are sparsely reported in the literature.

**Objectives:** This review aims to identify from the existing literature, those geometrical parameters that describe the three-dimensional anatomy of the talar neck.

**Methods:** A scoping literature review will be conducted in accordance with the PRISMA-ScR guidelines. The primary searches will be conducted on the PubMed, EMBASE and Scopus databases, using a pre-defined search strategy. Any original research study (whether clinical or cadaveric, prospective or retrospective, comparative or noncomparative), looking at the human talus neck geometry or morphology will be included. Studies that do not describe the talar neck geometry, animal studies, review articles, conference abstracts and case reports will be excluded. Geometrical parameters (including techniques to measure these) that describe the three-dimensional anatomy of the talar neck will be identified. Qualitative analysis will be done by means of appropriate tables and diagrams. If feasible, quantitative analysis to determine pooled estimated of geometrical parameters identified in the review will be conducted by a random-effects meta-analysis model. No formal risk-of-bias assessment has been planned, as this is a scoping literature review.

## 1. Background

The talus neck connects the talar head with the talar body. Fractures of the talar neck can alter this relationship. Comminution of the medial part of the talar neck is a common occurrence in talar fractures, and can result in a varus deformity if not addressed appropriately (1). Malunion after a talar neck fracture alters the talar head-body relationship and can thereby alter the biomechanics of the subtalar, talonavicular and ankle joints (2). Therefore, accurate assessment of talar neck geometry is vital to avoid malreduction and subsequent talar malunion (3). In established talar malunions, assessment of three-dimensional geometry is vital to plan operative correction.

## 2. Need for review

The literature on talar neck malunions is limited to description of the varus nature of the deformity and its surgical correction (4–6). The geometrical parameters that describe the three-dimensional anatomy of the talar neck are sparsely reported. Hence, this review aims to identify from the existing literature, those geometrical parameters that describe the three-dimensional anatomy of the talar neck.

## 3. Objectives

Primary Objectives:

a. To determine the geometrical parameters that describe the three-dimensional anatomy of the talar neck.
b. To determine the measurement techniques/methods for these parameters.

Secondary Objective

To determine the normative values of these parameters.

## 4. PICO framework for the study

a. Participants: human tali
b. Intervention: measurement of talar neck geometry by any method
c. Control: none
d. Outcomes: types, measurement techniques and values of different parameters that describe the talar neck geometry

## 5. Methods

This systematic review and meta-analysis will be conducted in accordance with the Preferred Reporting Items for Systematic Reviews and Meta-analysis – Scoping Review extension guidelines (PRISMA-ScR) (7).

a. *Review Protocol* A protocol of the review will be formulated a priori in accordance with the PRISMA-P guidelines. **(Appendix I)**
b. *Eligibility Criteria* Any original research study (whether clinical or cadaveric, prospective or retrospective, comparative or noncomparative), looking at the human talus neck geometry or morphology will be included. Both English and non-English articles will be included. Studies that do not describe the talar neck geometry, animal studies, review articles, conference abstracts and case reports will be excluded.
c. *Information Sources & Literature search* The primary search will be conducted on the PubMed, EMBASE and Scopus databases, using a pre-defined search strategy **(Table 1)**. For the secondary search, a manual search of references from the full-text of all included articles & relevant review articles will be conducted. There will be no restrictions on the language or date of publication.
d. *Study Selection* The titles and abstracts of all articles identified in the initial search will be screened independently by two authors. After initial screening, full-texts of the selected articles will be obtained. Eligibility for inclusion into the review will be assessed according to the pre-specified inclusion/exclusion criteria. Discrepancies will be resolved by mutual agreement. Reasons for exclusion of those studies for which full-text was obtained will be documented. A reference management software (Zotero) will be used to manage references.
e. *Data Collection & Data Items* Data will be extracted on pre-piloted data collection forms by two authors independently, a third author will check the data for accuracy. Baseline data items will include:
  - First author name, year and journal of publication
  - Language of publication
  - Study design: clinical/cadaveric; whether comparative or non-comparative; whether prospective or retrospective
  - Ethnicity
  - Number of subjects/bones evaluated
  - Number of males and females (if reported by the study)
  - Age distribution (if reported by the study)
  Once data extraction is complete, a spreadsheet containing the baseline characteristics, outcome measures and other important information, with respect to all studies included in the review will be created.
f. *Outcome Measures* Since this is a scoping review, it is not feasible to specify *all* outcome measures at the protocol stage. However, the following measures will be evaluated and additions and/or modifications will be made as needed:
  - Method(s) of measurement/evaluation
  - Geometrical parameter(s) evaluated: including, but not limited to talar inclination, declination, torsion, neck length etc.
  - Values of the geometrical parameter(s) evaluated (means and standard deviation)
g. *Data Analysis and Synthesis* Both qualitative and quantitative analyses will be performed. For qualitative analysis, appropriate tables and data visualization diagrams will be used. Baseline data items as well as all the pre-specified outcome measures will be reported. Quantitative analysis will be done to determine the pooled estimates of geometrical parameters if ≥ 2 studies describing the values of these parameters are included in the review. Pooled values of these parameters will be estimated by *random-effects* meta-analysis modelwill be reported as means with 95% confidence intervals. Forest plots will be constructed to visualize the results. Statistical heterogeneity will be evaluated by the *I*^2^ test. Leave-one-out sensitivity analysis will be performed if high statistical heterogeneity is identified (I^2^ > 75%). No subgroup analysis or meta-regression has been planned *a-priori*; however, it may be undertaken depending on the available evidence. Analysis will be performed by the *Open Meta Analyst* and *Stata MP version 14*.*0* software.
h. *Assessment of Risk of Bias* Since this is a scoping literature review, no formal risk-of-bias assessment has been planned.

**Table 1:** Search Strategy.

|  |  |
| --- | --- |
| **PubMed** | ((talus OR tali) OR (astragalus)) AND (((((((geometry OR (geometrical)) OR (geometric)) OR (morphometric)) OR (morphometry)) OR (measurement)) OR (biometry)) OR (three dimensional)) OR (three dimensional modeling)) **Filters:** Humans |
| **SCOPUS** | (( TITLE-ABS-KEY ( talus ) OR TITLE-ABS-KEY ( tali ) OR TITLE-ABS-KEY ( astragalus ) ) ) AND ( ( TITLE-ABS-KEY ( geometry ) OR TITLE-ABS-KEY ( morphometry ) OR TITLE-ABS-KEY ( measurement ) OR TITLE-ABS-KEY ( morphology ) OR TITLE-ABS-KEY ( biometry ) OR TITLE-ABS-KEY ( three AND dimensional AND modeling ) ) ) AND ( LIMIT-TO ( DOCTYPE , "ar" ) ) |
| **EMBASE** | (geometry OR morphometry OR measurement OR biometry OR 'three-dimensional imaging') AND (talus OR astragalus OR tali) |

## Data Availability

This is a protocol for a scoping literature review and as such, contains no study data. The search strategy for the protocol has been included in Table 1.

## APPENDIX 1

**PRISMA-P (Preferred Reporting Items for Systematic review and Meta-Analysis Protocols) 2015 checklist: recommended items to address in a systematic review protocol\***

*From: Shamseer L, Moher D, Clarke M, Ghersi D, Liberati A, Petticrew M, Shekelle P, Stewart L, PRISMA-P Group. Preferred reporting items for systematic review and meta-analysis protocols (PRISMA-P) 2015: elaboration and explanation. BMJ. 2015 Jan 2;349(jan02 1):g7647*.

| Section and topic | Item No | Checklist item | Page Number |
| --- | --- | --- | --- |
| **ADMINISTRATIVE INFORMATION** |  |  |  |
| Title: |  | Parameters for assessment of talar neck geometry: a protocol for scoping literature review | 1,2 |
| Identification | 1a | Identify the report as a protocol of a systematic review | 1,2 |
| Update | 1b | If the protocol is for an update of a previous systematic review, identify as such | NA |
| Registration | 2 | If registered, provide the name of the registry (such as PROSPERO) and registration number | NA |
| Authors: |  | Siddhartha Sharma, Karan Jindal, Sandeep Patel, Sharad Prabhakar, Mandeep S Dhillon. | 1 |
| Contact | 3a | Provide name, institutional affiliation, e-mail address of all protocol authors; provide physical mailing address of corresponding author | 1 |
| Contributions | 3b | Describe contributions of protocol authors and identify the guarantor of the review | 1 |
| Amendments | 4 | If the protocol represents an amendment of a previously completed or published protocol, identify as such and list changes; otherwise, state plan for documenting important protocol amendments | NA |
| Support: |  | None |  |
| Sources | 5a | Indicate sources of financial or other support for the review | 1 |
| Sponsor | 5b | Provide name for the review funder and/or sponsor – No sponsor | 1 |
| Role of sponsor or funder | 5c | Describe roles of funder(s), sponsor(s), and/or institution(s), if any, in developing the protocol | NA |
| **INTRODUCTION** |  |  |  |
| Rationale | 6 | Describe the rationale for the review in the context of what is already known | 3 |
| Objectives | 7 | Provide an explicit statement of the question(s) the review will address with reference to participants, interventions, comparators, and outcomes (PICO) | 3 |
| **METHODS** |  |  |  |
| Eligibility criteria | 8 | Specify the study characteristics (such as PICO, study design, setting, time frame) and report characteristics (such as years considered, language, publication status) to be used as criteria for eligibility for the review | 3, 4 |
| Information sources | 9 | Describe all intended information sources (such as electronic databases, contact with study authors, trial registers or other grey literature sources) with planned dates of coverage | 4 |
| Search strategy | 10 | Present draft of search strategy to be used for at least one electronic database, including planned limits, such that it could be repeated | Table 1, Page 8 |
| Study records: |  |  |  |
| Data management | 11a | Describe the mechanism(s) that will be used to manage records and data throughout the review | 5 |
| Selection process | 11b | State the process that will be used for selecting studies (such as two independent reviewers) through each phase of the review (that is, screening, eligibility and inclusion in meta-analysis) | 5 |
| Data collection process | 11c | Describe planned method of extracting data from reports (such as piloting forms, done independently, in duplicate), any processes for obtaining and confirming data from investigators | 5 |
| Data items | 12 | List and define all variables for which data will be sought (such as PICO items, funding sources), any pre-planned data assumptions and simplifications | 6 |
| Outcomes and prioritization | 13 | List and define all outcomes for which data will be sought, including prioritization of main and additional outcomes, with rationale | 6 |
| Risk of bias in individual studies | 14 | Describe anticipated methods for assessing risk of bias of individual studies, including whether this will be done at the outcome or study level, or both; state how this information will be used in data synthesis | Not planned |
| Data synthesis | 15a | Describe criteria under which study data will be quantitatively synthesized | 6 |
| | 15b | If data are appropriate for quantitative synthesis, describe planned summary measures, methods of handling data and methods of combining data from studies, including any planned exploration of consistency (such as $I^2$, Kendall's $\tau$) | 6 |
|  | 15c | Describe any proposed additional analyses (such as sensitivity or subgroup analyses, meta-regression) | Not planned |
|  | 15d | If quantitative synthesis is not appropriate, describe the type of summary planned | NA |
| Meta-bias(es) | 16 | Specify any planned assessment of meta-bias(es) (such as publication bias across studies, selective reporting within studies) | Not planned |
| Confidence in cumulative evidence | 17 | Describe how the strength of the body of evidence will be assessed (such as GRADE) | Not planned |

**\* It is strongly recommended that this checklist be read in conjunction with the PRISMA-P Explanation and Elaboration (cite when available) for important clarification on the items. Amendments to a review protocol should be tracked and dated. The copyright for PRISMA-P (including checklist) is held by the PRISMA-P Group and is distributed under a Creative Commons Attribution Licence 4.0.**

